# The effects of exercise and physical activity on gait biomechanics and pain in knee osteoarthritis: A systematic review and meta-analysis

**DOI:** 10.64898/2026.11.08.26367018

**Authors:** Nathan Bytheway, Shadi Amor, Paul Swinton, Kevin Cheah, Andrew Morrison, Hasmik Jasmine Samvelyan

## Abstract

**Objectives:** To evaluate the effects of exercise and physical activity on gait biomechanics and pain in adults with knee osteoarthritis.

**Methods:** Systematic searches were performed of PubMed, Web of Science, Scopus and SPORTDiscus databases for controlled intervention studies published up to 31 December 2024. Eligible studies included adults with knee osteoarthritis undertaking exercise or physical activity compared with control group(s). Risk of bias was assessed using the Downs and Black Checklist. Bayesian three-level hierarchical random-effects meta-analyses synthesised standardised mean differences (SMD; effect size) with 95% credible intervals (CrIs).

**Results:** Of 2,602 records identified, 18 studies involving 1,102 participants (345 males and 757 females) were included. Exercise and physical activity reduced pain (SMD 0.61, 95% CrI 0.29 to 0.92) and improved spatiotemporal gait parameters (SMD 0.67, 95% CrI 0.27 to 1.05). Improvements were reported across several intervention types, including strengthening, aerobic, multicomponent, and mind–body exercise. Effects on knee kinematics (SMD 0.12, 95% CrI −0.18 to 0.41) and kinetics (SMD −0.03, 95% CrI −0.25 to 0.18) were small or uncertain.

**Conclusions:** Exercise and physical activity may improve pain and spatiotemporal gait characteristics in knee osteoarthritis, but consistent improvements in knee kinematics and kinetics were not demonstrated. Further adequately powered trials with standardised biomechanical assessments and longer assessments are needed.

*Registration:* PROSPERO CRD42024565957.

**Key messages:** *What is already known on this topic:* Exercise and physical activity are recommended non-pharmacological interventions for managing knee osteoarthritis, but their effects on gait biomechanics across different exercise modalities remain unclear.

*What this study adds:* This systematic review and meta-analysis found that exercise and physical activity were associated with reduced pain and improved spatiotemporal gait parameters in people with knee osteoarthritis, while effects on knee kinematics and kinetics were small or uncertain.

*How this study might affect research, practice or policy:* Exercise/physical activity remain central to knee osteoarthritis management. Future trials should investigate whether targeted interventions can modify knee joint mechanics, using standardised biomechanical assessments and longer follow-up.

## Introduction

Knee osteoarthritis, a chronic musculoskeletal disease, is the most prevalent type of osteoarthritis and the leading cause of disability and major healthcare costs worldwide. Globally, approximately 13% of women and 10% of men aged over 60 years have symptomatic knee osteoarthritis. The prevalence of knee osteoarthritis is expected to continue to increase globally due to ageing population and increasing rates of obesity and injury [1].

Knee osteoarthritis is a whole joint disease involving multifaceted interactions between structural, biological, and biomechanical factors. It is characterised by a progressive deterioration of the articular cartilage, formation of osteophytes, subchondral bone sclerosis, synovial proliferation, inflammation, and lax tendons. These ultimately lead to a loss of knee joint function, chronic pain, impaired mobility and proprioception, and disability.

Management of symptoms and pain are primary treatments for knee osteoarthritis [2], with knee replacement surgery typically considered later in the disease. While medications such as non-steroidal anti-inflammatory drugs may be prescribed to relieve pain, physical activity and exercise are non-pharmacological recommendations for a long-term management of knee osteoarthritis [3].

Patients with knee osteoarthritis exhibit altered gait biomechanics when compared to age-matched healthy individuals [4, 5]. Common gait changes include spatiotemporal parameters such as reduced walking speed and stance duration as well as increased step length. Three-dimensional patterns of the knee joint such as kinematic changes include reduced range of motion in knee flexion, extension and rotation, and increased knee adduction, whereas kinetic changes include reduced peak knee extension moment, increased knee valgus moment and loading on the contralateral limb. Moreover, alterations in gait biomechanics can impact other joints increasing likelihood of joint tissue damage, muscular injury and chronic pain.

Increasing evidence suggests that physical activity and exercise training are cost-effective and widely available interventions for the treatment of knee osteoarthritis, however, consideration of type of physical activity and optimal exercise modality are needed if the aim is to reduce pain, improve gait, knee joint function, stiffness, and muscle weakness in individuals with knee osteoarthritis.

For example, low-impact physical activities including walking, swimming, and cycling have been shown to be beneficial for individuals with severe knee osteoarthritis [6]. Regular aquatic exercises can help to relieve pain and improve daily function in people with both knee and hip osteoarthritis [7, 8], whereas high-impact activities such as running and jumping can potentially worsen symptoms of knee osteoarthritis by increasing stress and inflammation in affected joints [9, 10].

In this systematic review with meta-analysis, we examine the effects of different types of exercise training and physical activity on gait characteristics and pain in individuals with knee osteoarthritis and provide evidence to inform personalised therapeutic strategies as well as a reference for future research and clinical application of these training programmes.

## Methods

### Study eligibility criteria

Potential studies were searched and screened according to their population, intervention, comparator, outcome, and study design (PICOS) (Table 1). Studies not meeting the criteria outlined in Table 1 were excluded.

**Table 1.** Study eligibility criteria.

| **Criteria** |
|---|
| 1) Study purpose focused on knee osteoarthritis and evaluation of an intervention with a therapeutic effect |
| 2) Published in English |
| 3) Adults with knee osteoarthritis aged 18 years and older from any race, gender or ethnicity |
| 4) Physical activity or exercise intervention compared with either a control group or a SHAM (inactive intervention) |
| 5) Data available for changes in gait parameters and/or pain |

#### Population

Adults aged 18 years and older from any race, gender, ethnicity, or geographical location. Studies conducted on populations with knee osteoarthritis and comorbidities that severely affected gait at baseline (such as individuals diagnosed with inflammatory arthritis, neurological conditions including Parkinson’s disease) were excluded. Studies that assessed exercise/physical activates following surgical intervention were also excluded.

#### Intervention

Any physical activity or exercise intervention that involved motion of the lower limbs aiming to improve gait characteristics, increasing mobility, and reducing or relieving pain. The characteristics of the physical activity/exercise regimen (such as type, duration, total work) were unrestricted but were considered during interpretation of results. Any study that insufficiently described the intervention was excluded.

#### Comparator

Only studies of physical activity or exercise interventions that included a control group of participants with established knee osteoarthritis were included. Alternate interventions were excluded unless a control group provided relevant information based on the inclusion and exclusion criteria. Studies without a control or comparator group were excluded.

#### Outcomes

The primary outcomes were gait related characteristics including spatiotemporal (such as gait velocity and cadence, step length and width), kinematic (such as range of motion (ROM), knee flexion and extension) or kinetic (such as joint moment and ground reaction force (GRF)) parameters used to assess gait patterns [4, 11].

All included studies reported gait biomechanics such as spatiotemporal, kinetic, and kinematic parameters. Lower limb biomechanics were extracted from studies that used marker-based motion capture or two-dimensional (2D) video capture from applications such as Kinovea, Vicon or any other method of capturing biomechanical gait parameters. Studies investigating static alignment or non-gait biomechanical metrics were excluded.

The secondary outcomes included assessment of pain and mobility following physical activity/exercise intervention through commonly used questionnaires including Western Ontario and McMaster Universities Arthritis Index (WOMAC), knee osteoarthritis outcome score (KOOS), visual analogue scale (VAS), or any other validated pain assessment tool [12–14].

#### Study design

Controlled intervention studies were included including randomised clinical trials and quasi-experimental designs. Studies that implemented a secondary analysis of data were also considered if the study details were outlined.

Conference proceedings, abstracts, case studies, dissertations, intervention studies with no observation/control group, and reviews (systematic or narrative) were excluded.

This meta-analysis adhered to the Preferred Reporting Items for Systematic reviews and Meta-Analyses (PRISMA) guidelines [15] (Tables S1 and S2). The protocol for this meta-analysis was preregistered in PROSPERO (CRD42024565957) [16].

## Data sources and search strategy

We searched four electronic databases: PubMed, Web of Science, Scopus, and Sport Discus from 13 July 2024 to 31 December 2024. Key search terms included ‘Osteoarthritis’, ‘Knee’, ‘Gait’, AND (‘Exercise OR physical activity’). Based on PRISMA guidelines [15], an example of the search strategy is provided in the Supplementary material (Table S3).

Each database was searched using tailored syntax and relevant MeSH terms, adjusted accordingly for each database. Where appropriate, headline headings were also used if the search string resulted in a greater number of returned results. Only studies involving human participants were considered. All selected articles were written in English (Table S4).

The first author (NB) conducted all electronic database searches. In addition, cross-referencing of retrieved studies was also conducted to ensure completeness of the search.

## Study records and selection

All studies were imported into RefWorks reference manager (RefWorks v.3, Clarivate, USA). A tsv (tab separated values) file was created to import all studies into Microsoft Excel, and duplicates removed electronically and manually by the first author (NB). A copy of the reference database was then provided to the second author (SA) for dual screening. Screening was conducted independently by the two authors (NB and SA). Reviewers were not blinded to the journal titles or study authors/affiliations. Reasons for exclusion were coded based on one or more of the following: (1) inappropriate population, (2) inappropriate intervention, (3) inappropriate comparison(s), 4) inappropriate outcome(s), (5) inappropriate study design or (6) other. On completion, the screeners met to discuss their selections and reconciled any discrepancies by consensus. If an agreement could not be achieved, the fifth author (AM) provided a recommendation. The precision of searches was calculated as the number of studies included divided by the number of studies screened (less duplicates), we then calculated the number-needed-to-screen (NNS) by taking the reciprocal of the precision [17].

## Data extraction and coding

Prior to data extraction, an electronic codebook was developed (Table S5). Data were then independently extracted into a spreadsheet.

The extracted data were coded based on the following major categories: (1) study characteristics (e.g., author, year, title, journal, funding and conflict of interest statement, aim, study design overview, etc.); (2) participant characteristics (e.g. number, sex, age, height, mass, body mass index, etc.), (3) intervention details (e.g., the exercise or physical activity type, duration, components, frequency, etc.), and (4) outcome characteristics (e.g., biomechanical gait parameters categorised into spatiotemporal, kinematic, and kinetic parameters, along with pain scales used in each study, means and SDs, CIs etc.). Other data points extracted included study length and whether exercise or physical activity produced a positive outcome relative to the comparator group.

Coding details are available in the accompanying codebook (Table S5). Two reviewers (NB and SA) extracted data independently before meeting to resolve any discrepancies by consensus. If an agreement could not be achieved, the third reviewer (AM) provided a recommendation.

## Risk of bias assessment

Risk of bias and the quality of studies were assessed using The Downs and Black Checklist [18]. All domains of the checklist were assessed for each article including reporting, external validity, internal validity (bias and confounding variables (selection bias)), and power.

Two reviewers (NB and SA) undertook the risk of bias assessment independently before meeting to resolve any discrepancies by consensus. Where this could not be achieved, the third reviewer (AM) provided a recommendation.

## Data synthesis

Meta-analysis was performed by each subgroup of biomechanical gait variables that was extracted from each study. Extracted data were transformed into pairwise-controlled (between-group) standardised mean difference effect sizes. This was achieved by subtracting the comparator mean difference from the intervention mean difference and dividing by the pooled baseline standard deviation. Standard distributional assumptions were used to calculate effect size standard errors.

In the present meta-analysis, a Bayesian framework was implemented through three-level hierarchical models with random effects to account for variation in study mean effect, and three-level models included to model covariance of multiple outcomes reported in the same study [19].

Standard distributional assumptions for effect size standard errors based on pre- and post-intervention data require an estimate of their correlation, which is rarely reported in studies. To account for this uncertainty, a standard value of 0.7 was used to generate an informative prior for the correlation, with variance included to account for correlations ranging from 0.5 to 0.9. Predictive distributions developed for meta-analyses were obtained and transformed into gamma distributions. Inferences were performed on posterior samples generated by Hamiltonian Markov Chain Monte Carlo, with Bayesian 95% and 75% credible intervals (CrIs) used for location and variance parameters, respectively.

Interpretations were based on visual inspection of the posterior sample, the median value (SMD_0.5_: 0.5-quantile), CrIs, and the probability that the pooled mean exceeded small (0.2), medium (0.5), and large (0.8) thresholds. Analyses were performed using the R wrapper package brms, which interfaced with Stan to perform sampling. Convergence of parameter estimates was confirmed for all models with Gelman-Rubin R-hat values below 1.1.

## Results

### Study characteristics

The search yielded in 2602 records, of which 83 studies were retained for full screening and 18 studies met the eligibility criteria for final inclusion (Figure 1) comprising of 1,102 participants (345 males and 757 females).

**Figure 1.**
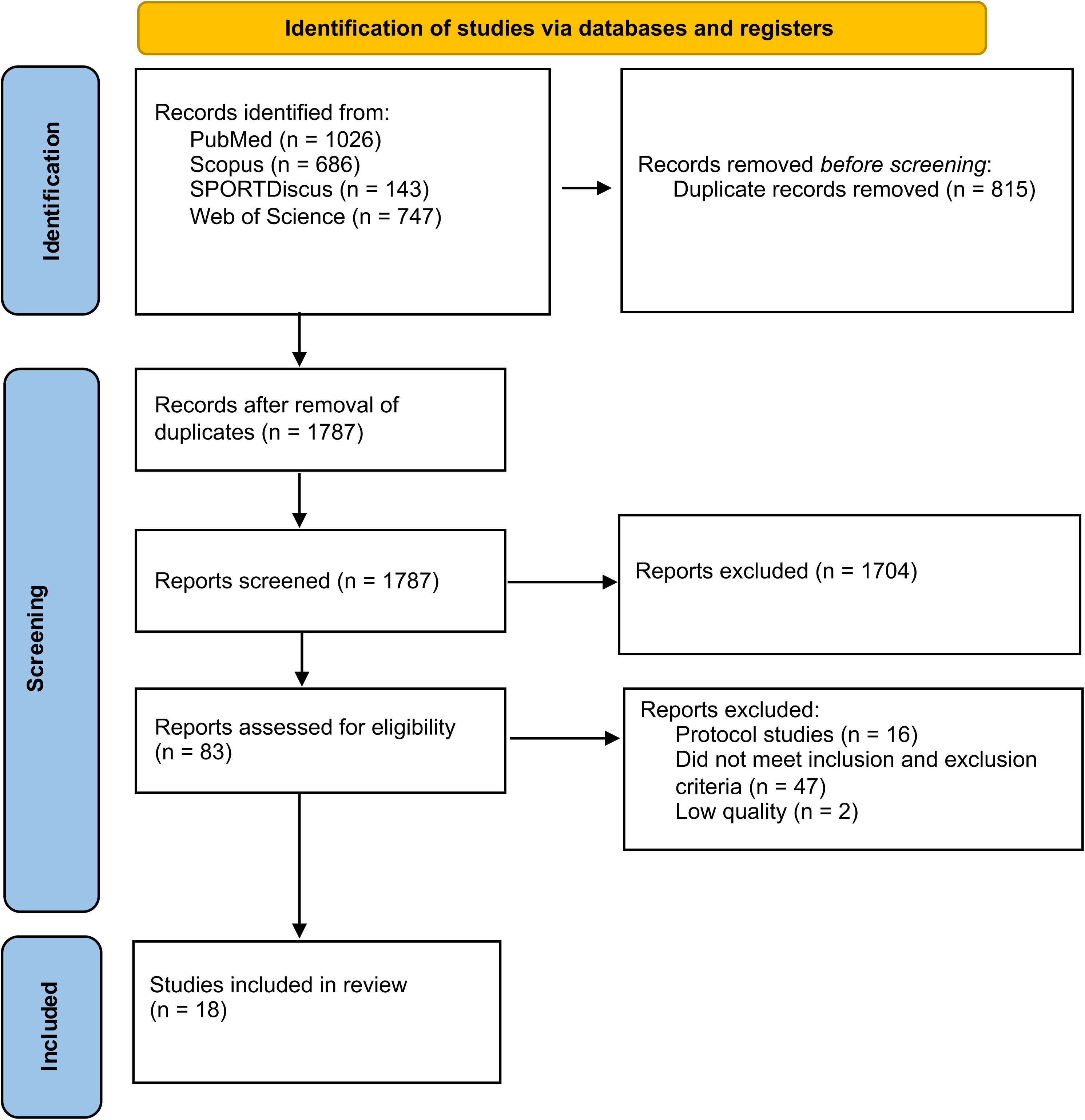
Search flow diagram

A flow diagram that elicits the search and selection processes is shown in Figure 1. A full list of studies is available in Table S6.

General description of the studies included can be found in Table 2. All included studies were published in peer-reviewed English-language journals, starting in 1997 and ending in 2023. The studies were conducted in a variety of countries; six in Australia, three in USA and India, two in Iran and China, and one each in Canada and Japan.

**Table 2.** Study characteristics.

| Study | Country | Sample size (%F) | Age Mean (SD) - unless otherwise stated | BMI Mean (SD) - unless otherwise stated | Participants | Exercise/physical activity intervention | Gait assessment | Knee pain assessment | Baseline OA severity (KL) % - unless otherwise stated |
|---|---|---|---|---|---|---|---|---|---|
| Aoki et al., 2009<sup>31</sup> | Japan | 36 (100) | **Control**<br>74.4 (6.4)<br>**Intervention**<br>72.3 (5.2) | 26.2 (2.6) | Home-based, participants with severe knee osteoarthritis (KOA) randomly allocated to stretching (n = 17) or control (n = 19) group, age > 70 years | **Stretching**<br>Home-based knee-flexion stretching exercises performed daily for approximately 80 days. | Three-dimensional optical motion capture (Mac3D; six cameras) with two force plates; sampling frequency 60 Hz. | 100-mm visual analogue scale (VAS),<br>> 4/10 | **Control**<br>Grade<br>IV (IV to IV)<br>**Intervention**<br>Grade<br>IV (III to IV) |
| Azizi et al., 2020<sup>26</sup> | Iran | 32 (0) | **Control**<br>65.5 (3.3)<br>**Intervention**<br>63.5 (4.7) | > 34 kg/m<sup>2</sup> | Community pool, with KOA randomly allocated to aquatic exercise (n = 16) or control (n = 16) group, age ≥ 60 years | **Aquatic**<br>Supervised pool-based exercise three times weekly for 60 minutes per session, incorporating warm-up, strengthening and cool-down exercises, for 8 weeks. | High-speed video camera (Casio FH20 with software Kinovea software for gait image analysis. | 100-mm VAS<br>> 5/10 | Not reported |
| Bennell et al., 2010<sup>22</sup> | Australia | 89 (48) | **Control**<br>64.6 (7.6)<br>**Intervention**<br>64.5 (9.1) | 28 (4.4) | Home exercise, with KOA randomly allocated to exercise (n = 45) or control (n = 44) group, age > 50 years, BMI < 35 kg/m<sup>2</sup> | **Strengthening**<br>Home-based hip abductor and adductor strengthening exercises, supplemented by seven physiotherapy sessions over 12 weeks; performed five times per week. | Three-dimensional motion analysis using an eight-camera Vicon system (120 Hz) and the Plug-in-Gait marker set. | Western Ontario and McMaster Universities Osteoarthritis Index (WOMAC) | **Control**<br>Grade<br>II = 34.1<br>III = 31.8<br>IV = 34.1<br>**Intervention**<br>Grade<br>II = 33.3<br>III = 33.3<br>IV = 33.3 |
| Bhore and Shinde, 2023<sup>35</sup> | India | 120 (68) | 57.7 (7.7) | 29.2 (6.4) | With bilateral KOA randomly allocated to multicomponent exercise (n = 60) and conventional control group (n = 60), aged 50 - 65 years, KL 1 and 3 | **Multi-component**<br>Programme comprising core and lower-limb strengthening, proprioception and balance exercises, performed five times weekly for 45–60 minutes per session over 6 weeks. | Not reported. | WOMAC | Not reported |
| Chen et al., 2021<sup>28</sup> | China | 18 (72) | 59 (8.3) | 23.1 (1.3) | Recruited from the Department of Rehabilitation Medicine at the Fifth Affiliated Hospital of Guangzhou Medical University with mild-to-moderate KOA, randomly allocated to exercise group with lower body pressure treadmill (n = 8, %F = 75) and conventional control group (n = 10; %F =70), aged 50 - 75 years | **Aerobic**<br>Lower-body positive-pressure treadmill walking six days weekly for 30 minutes per session over 2 weeks, including 20 minutes of walking at 65% body weight, with warm-up and cool-down periods. | Three-dimensional motion analysis using the BTS Smart DX 7000 system, with 22 reflective markers positioned according to the Davis protocol. | WOMAC | **Control**<br>Grade<br>II = 72.2<br>Grade<br>III = 28<br>**Intervention**<br>Grade<br>II = 75<br>Grade<br>III = 100 |
| Fransen et al., 2001<sup>30</sup> | Australia | 126 (73) | **Control**<br>66.1 (10.3)<br>**Intervention**<br>Individual<br>68.5 (8.7)<br>Group<br>65.3 (7.1) | **Control**<br>28.3 (4.5)<br>**Intervention**<br>Individual<br>30.0 (4.6)<br>Group<br>29.9 (5.9) | Participants with KOA had knee pain on most days of the past month, and evidence of radiographic disease were randomly allocated to 3 groups: individual treatments (n = 43), group format program (n = 40) and waiting-list control group (n = 43), age > 50 years | **Strengthening**<br>Individualised treatment: Prescribed by a physiotherapist.<br>Group exercise: Supervised sessions twice weekly for 1 hour, supplemented by home exercise three times weekly, incorporating stretching, stationary cycling and lower-limb strengthening for 8 weeks. | Eight-metre electronic footswitch walkway. | WOMAC | Not reported |
| Foroughi et al., 2011<sup>33</sup> | Australia | 54 (100) | **Control**<br>65 (7)<br>**Intervention**<br>66 (8) | **Control**<br>32.7 (8.4)<br>**Intervention**<br>31.4 (5.4) | Conducted at the University of Sydney under supervision of exercise physiologists, participants with KOA at least in one knee were randomly allocated to progressive resistance training exercise (n = 26) and sham-exercise control group (n = 28), age > 40 years | **Resistance**<br>Supervised, high-intensity progressive resistance training (80% of peak muscle strength), three times weekly for 6 months, targeting the lower-limb muscles. | Three-dimensional motion analysis using 10 Eagle cameras (Motion Analysis Corporation), EvaRT software and two sets of eight-channel force plates. | WOMAC | Modified Outerbridge Classification (Range 1-4; Mild, Moderate, Severe, Very severe)<br>**Control**<br>Grade<br>I = 39.3<br>II = 7.1<br>III = 35.7<br>IV = 7.1<br>**Intervention**<br>Grade<br>I = 34.6<br>II = 19.2<br>III = 38.5<br>IV = 3.8 |
| Foroughi et al., 2011<sup>23</sup> | Australia | 54 (100) | 64 (7) | 32.6 (6.9) | Conducted at the University of Sydney under supervision of exercise physiologists, participants with KOA at least in one knee were randomly allocated to supervised progressive resistance training exercise group (n = 26) or sham-exercise control group (n = 28), age > 40 years | **Resistance**<br>Supervised, high-intensity progressive resistance training (80% of peak muscle strength), three times per week for 6 months, targeting the lower-limb muscles. | Three-dimensional motion analysis using 10 Eagle cameras (Motion Analysis Corporation), EvaRT software and two sets of eight-channel force plates. | WOMAC | Modified Outerbridge Classification (Range 1-4; Mild, Moderate, Severe, Very severe)<br>**Control**<br>Grade<br>I = 42.9<br>II = 10.7<br>III = 25.0<br>IV = 21.4<br>**Intervention**<br>Grade<br>I = 23.1<br>II = 19.2<br>III = 19.2<br>IV = 38.5 |
| Joshi, Singh and Vij, 2019<sup>32</sup> | India | **Control**<br>21 (61.9)<br>**Intervention**<br>21 (42.9) | **Control**<br>54.3 (10.2)<br>**Intervention**<br>50.7 (7.3) | **Control**<br>28.1 (8.9)<br>**Intervention**<br>28.6 (10.2) | Conducted at the Department of Physiotherapy, Guru Jambheshwar University of Science and Technology.<br>Participants with KOA were randomly allocated to retro walking group<br>and conventional exercise control group, age > 40 years, KL < 4 | **Multi-component and aerobic**<br>Conventional physiotherapy exercises, including range-of-motion, strengthening and stretching, with the intervention group additionally undertaking treadmill retro-walking for 10 minutes per session, three times weekly for 6 weeks. | Footprint method and 10-metre walkway. | VAS | Not reported |
| Kean et al., 2017<sup>37</sup> | Australia | **Control**<br>48 (56)<br>**Intervention**<br>49 (55) | **Control**<br>63.8 (8.2)<br>**Intervention**<br>65.7 (8.2) | **Control**<br>28.5 (6.7)<br>**Intervention**<br>28.6 (4.4) | Secondary analysis of a 12-week RCT that evaluated the effects of quadriceps strengthening on the knee adduction moment, pain and physical function in people with knee osteoarthritis, age > 60 years, KL 2-4<br>The sample overlaps with the original trial reported by Lim *et al.*, 2008. | **Strengthening**<br>Home-based quadriceps strengthening programme comprising five non-weight-bearing exercises, performed five times weekly for 12 weeks. | Three-dimensional motion analysis using an eight-camera Vicon system, the Plug-in-Gait marker set and two force plates. | Not reported | **Control**<br>Grade<br>II = 29<br>III = 27<br>IV = 44<br>**Intervention**<br>Grade<br>II = 28.5<br>III = 28.5<br>IV = 43.0 |
| Lim et al., 2008<sup>21</sup> | Australia | **Control**<br>More neutrally aligned<br>28 (61)<br>More malaligned<br>26 (46)<br>**Intervention**<br>More neutrally aligned<br>27 (63)<br>More malaligned<br>26 (50) | **Control**<br>More neutrally aligned<br>60.8 (7.8)<br>More malaligned<br>66.6 (8.9)<br>**Intervention**<br>More neutrally aligned<br>64.1 (9.3)<br>More malaligned<br>67.2 (6.7) | **Control**<br>More neutrally aligned<br>28.4 (5)<br>More malaligned<br>30.3 (5.3)<br>**Intervention**<br>More neutrally aligned<br>29.0 (5.2)<br>More malaligned<br>28.2 (3.7) | Participants with tibiofemoral joint KOA in at least one knee, with medial knee pain, medial compartment osteophytes, and medial joint space narrowing greater than lateral joint space narrowing.<br>Participants with ≥ 5 degrees varus malalignment were deemed more malaligned (n = 52), and those with < 5 degrees malalignment (in either direction) were deemed more neutral (n = 55).<br>Each group was then randomised to receive either quadriceps strengthening intervention or no intervention (control) group, KL 2-4 | **Strengthening**<br>Home-based quadriceps strengthening programme, performed five times weekly for 12 weeks, with progressive increases in exercise volume. | Three-dimensional motion analysis using an eight-camera Vicon system, the Plug-in-Gait marker set and two force plates. | WOMAC | **Control**<br>More neutrally aligned<br>Grade<br>II = 54<br>III = 36<br>IV = 11<br>More malaligned<br>Grade<br>II = 12<br>III = 15<br>IV = 73<br>**Intervention**<br>More neutrally aligned<br>Grade<br>II = 44<br>III = 26<br>IV = 30<br>More malaligned<br>Grade<br>II = 15<br>III = 31<br>IV = 54 |
| Messier, Thomson and Ettinger, 1997<sup>20</sup> | USA | 103 (74) | **Control**<br>69.2 (1)<br>**Intervention**<br>Aerobic<br>70.3 (1.3)<br>Weight training<br>67.2 (0.9) | **Control**<br>32.5 (0.9)<br>**Intervention**<br>Aerobic<br>31.4 (1)<br>Weight training<br>30.1(0.9) | Participants with radiographic evidence of KOA randomised to aerobic (n = 33), weight training (n = 34) or health education (control) group (n = 36), age ≥ 60 years | **Resistance and aerobic**<br>Aerobic training: Walking programme, three times weekly, incorporating warm-up, 40 minutes of walking at 50–80% heart-rate reserve and cool-down.<br>Weight training: Supervised resistance training involving upper- and lower-body exercises, three times weekly.<br>Both programmes lasted 18 months, with 3 months of facility-based and 15 months of home-based training. | Photoelectric timing system (Lafayette Model 63501), VP320 video camera system and walkway incorporating an AMTI force platform. | Scale specifically developed for patients with knee osteoarthritis | Not reported. |
| Paul et al., 2016<sup>24</sup> | India | 40 (55) | **Control**<br>54.2 (5.8)<br>**Intervention**<br>53.1(6.3) | **Control**<br>25.7 (2.9)<br>**Intervention**<br>25.8 (1.6) | Interventions were conducted at the department of Physiotherapy, National Institute for the Orthopedically Handicapped, Kolkata, WB, India. Participants with unilateral KOA randomised to receive either hip and knee strengthening exercise (n = 20) or knee muscle strengthening exercise (control) (n = 20), age > 50 years, KL 1-3 | **Strengthening**<br>Supervised hip and knee muscle strengthening compared with knee-only strengthening, performed three times weekly for 45 minutes per session over 6 weeks. | Computer Dynography (CDG; Infrotronics Medical Industrial Engineering). | The reduced WOMAC and Numeric Rating Scale (NRS) | Not reported. |
| Salacinski et al., 2012<sup>34</sup> | USA | 37 (73) | 57.7 (9.8) | 23.8 (5.3) | Participants were randomly assigned to intervention (n = 19) or control group (n = 18), aged 37 - 74 years, KL 1-3 | **Aerobic**<br>Community-based group cycling, with at least two supervised sessions weekly. Sessions progressed from 40 to 60 minutes and included cycling intervals and simulated hill climbs, over 12 weeks. | GAITRite instrumented walkway (3.66 m; CIR Systems). | Likert-scale WOMAC<br>Knee Injury and Osteoarthritis Outcome Score (KOOS)<br>Visual Analog Pain Scale (VAPS) | Not reported |
| Sedaghatnezhad et al., 2021<sup>29</sup> | Iran | **Control**<br>15 (73)<br>**Intervention**<br>15 (93) | **Control**<br>59.6 (7.43)<br>**Intervention**<br>53.8 (7.43) | **Control**<br>27.7 (1.93)<br>**Intervention**<br>26.55 (2.08) | Participants with KOA were randomly assigned to physical therapy plus uphill treadmill walking (n = 15) or physical therapy (control) group (n=15), aged 45 - 60 years,<br>KL 2-3 | **Aerobic**<br>Uphill treadmill walking (8° incline) at 1.1 m/s for 30 minutes per session, five days weekly for 2 weeks, in addition to routine physiotherapy. | 10-metre walking speed test: self-selected walking speed measured over the middle 10 m of a 20-m walkway using a stopwatch; three trials, with mean time used to calculate walking speed. | VAS | **Control**<br>Grade<br>II= 40<br>III = 60<br>**Intervention**<br>Grade<br>II = 60<br>III = 40 |
| Sled et al., 2010<sup>36</sup> | Canada | **Control**<br>40 (56)<br>**Intervention**<br>40 (56) | **Control**<br>64.13 (9.04)<br>**Intervention**<br>62.98 (9.73) | **Control**<br>24.04 (3.24)<br>**Intervention**<br>27.38 (5.47) | Participants with medial KOA were allocated to home strengthening program for the hip abductor muscles (n=40) or control group (n=40), aged<br>> 40 years, KL 1-3 | **Strengthening**<br>Home-based hip abductor strengthening programme, including progressive resistance exercises and single-leg stabilisation activities, performed 3–4 times weekly for 8 weeks. | Optoelectronic motion tracking using two Optotrak 3020 cameras and two AMTI force plates along an 8-metre walkway. | WOMAC | Not reported |
| Vincent and Vincent, 2020<sup>27</sup> | USA | 88 (67.4) | 68.3 (6.4) | 30.4 (6.9) | Participants from Orthopaedic Clinics KOA with tibiofemoral KOA randomised to either concentrically focused resistance training (CNCRT) (n = 28) or eccentrically focused resistance training (ECCRT) (n = 30) or control group (n = 29), aged 60 -85 years,<br>KL 2 or 3 | **Resistance**<br>Concentric- or eccentric-focused resistance training involving upper- and lower-body exercises, performed twice weekly for 4 months. | GAITRite instrumented gait mat (8 m); walking at a self-selected speed. | NRS | Not reported |
| Zhu et al., 2016<sup>25</sup> | China | 46 (100) | **Control**<br>64.53 (3.43)<br>**Intervention**<br>64.61(3.40) | **Control**<br>25.05(3.42)<br>**Intervention**<br>25.23( 3.46) | In laboratory at the Shanghai University of Sport participants with KOA randomly allocated to Tai Ji Quan (n = 23) or control (attended one 60 min wellness education lecture bi-weekly) group (n = 23), aged 60 - 70 years, KL 1-3 | **Mind-body**<br>Tai Ji Quan training incorporating eight forms, performed three times weekly for 60 minutes per session over 24 weeks. | Sixteen-camera infrared motion analysis system (Vicon; Oxford Metrics). | WOMAC | **Control**<br>Grade<br>I = 26<br>II = 61<br>III = 13<br>**Intervention**<br>Grade<br>I = 30<br>II = 52<br>III = 17 |

Sixteen studies were randomised controlled trials (RCTs) either single-blind [20–29] or non-open-label [30–35].

One clinical trial used a quasi-experimental (non-equivalent pretest-posttest control group) design [36], and one study [37] was a secondary exploratory analysis from the previous RCT study [21].

One study [38] did not include all data in the manuscript and following emails to the corresponding author there was no response.

### Participant characteristics

Baseline participant characteristics of included studies are reported in Table 2.

#### Exercise/physical activity intervention characteristics

A description of the exercise/physical activity interventions are included in the Table 2. Of these included studies six studies investigated strengthening (hip and knee joint muscles) [21, 22, 24, 30, 36, 37], four resistance training [20, 23, 27, 33], five aerobic; four walking [20, 28, 29, 32] and one cycling [34], two multi-component [32, 35], one aquatic [26], one stretching [31] and one mind-body (Tai Ji Quan) [25] exercise/physical activity interventions as therapeutic strategies to improve gait characteristics (outcome 1) and reduce/relieve pain (outcome 2). The number of interventions exceeded the number of studies as two studies had more than one type of intervention [20, 32].

#### Assessment characteristics

##### Gait assessment characteristics

Thirteen studies assessed spatiotemporal gait parameters [20, 24–35]. Six studies assessed kinetic gait parameters [21–23, 33, 36, 37] and five studies assessed kinematic gait parameters [22, 23, 25, 31, 33].

Three studies [21, 22, 25] and one follow up study [37] used 3D motion analysis system (Vicon, Oxford Metrics, Oxford, UK) with cameras, two studies used three-dimensional motion measurement system (Motion Analysis Corporation, USA) with 10 Eagle video cameras, the Expert Vision Advanced software package (EvaRT5.0) and 2 sets of eight channel force plates (Kistler, Switzerland) [23, 33], one study used three-dimensional optical motion capture system with 6 cameras (Mac3D system; Motion Analysis Co., CA) and two force plates [31], while another study used Casio FH20 high-speed camera with Kinovea 0.8.15 software [26]. One study used BTS Smart DX 7000 (Bioengineering Technology System, Milan, Italy) [28], one study used CDG (Computer Dynography) (Infrotronics Medical Industrial Engineering; Netherlands) [24] whilst another study used Lafayette Model 63501 photoelectric control system with VP320 video camera system and walkway with AMTI force platform [20].

Two studies used either walkway [32] or walking speed test [29] (each 10 meter). One study used a walkway with two Optotrak 3020 optoelectronic motion tracking cameras along with two AMTI forceplates [36], other studies used either 12 ft (3.66 meter) GAITRite instrumented (CIR Systems, Inc, Sparta, NJ) [34] or electric footswitch walkways [30] and gait mat at a self-selected speed (GAITRite; CIR Systems Inc, Havertown, PA) [27] (all 8m long).

One study did not report the measurement system used for gait assessment [35].

##### Pain assessment characteristics

Apart from one study which used a scale specifically developed for their patients to assess the frequency and intensity of pain [20], all of the studies assessed pain using validated questionnaires including WOMAC, KOOS or numeric pain rating scale such as VAS or NRS.

Twelve studies used WOMAC [21–25, 28, 30, 33–37], whilst five studies used VAS for assessment of pain [26, 28, 29, 31, 32]. Two studies reported pain using NRS scale [24, 27] and only one study considered KOOS questionnaire [34]. Three studies used a combination of pain assessment tools [24, 28, 34].

Some studies included participants with mild (Kellgren-Lawrence grades KL 1-2), others severe (KL 3-4) knee osteoarthritis.

#### Risk of bias assessment

Methodological quality was assessed using the original Downs and Black checklist, with a maximum possible score of 32 points. Assessment using the Downs and Black Checklist is shown in Table 3.

**Table 3.** Risk of bias assessment.

| **Study** | **Reporting** | **External validity** | **Internal validity - bias** | **Internal validity - confounding** | **Power** | **Total score (out of 32)** |
|---|---|---|---|---|---|---|
| Aoki *et al.*, 2009<sup>31</sup> | 9 | 0 | 5 | 2 | 5 | 21 |
| Azizi *et al.*, 2020<sup>26</sup> | 10 | 1 | 4 | 4 | 5 | 24 |
| Bennell *et al.*, 2010<sup>22</sup> | 10 | 1 | 6 | 6 | 5 | 28 |
| Bhore and Shinde, 2023<sup>35</sup> | 7 | 0 | 4 | 3 | 5 | 19 |
| Chen *et al.*, 2021<sup>28</sup> | 10 | 1 | 5 | 4 | 5 | 25 |
| Foroughi, Smith, Lange, Baker, *et al.*, 2011<sup>33</sup> | 10 | 1 | 6 | 5 | 5 | 27 |
| Foroughi, Smith, Lange, Singh, *et al.*, 2011<sup>23</sup> | 11 | 1 | 6 | 5 | 4 | 27 |
| Fransen, Crosbie, Edmonds, 2000<sup>30</sup> | 10 | 2 | 6 | 6 | 5 | 29 |
| Joshi, Singh, Vij, 2019<sup>32</sup> | 7 | 0 | 4 | 3 | 5 | 19 |
| Kean *et al.*, 2017<sup>37</sup> | 9 | 1 | 6 | 5 | 5 | 26 |
| Lim *et al.*, 2008<sup>21</sup> | 11 | 1 | 6 | 6 | 5 | 29 |
| Messier, Thomson and Ettinger, 1997<sup>20</sup> | 8 | 1 | 5 | 5 | 5 | 24 |
| Paul *et al.*, 2016<sup>24</sup> | 9 | 1 | 6 | 4 | 5 | 25 |
| Salacinski *et al.*, 2012<sup>34</sup> | 11 | 1 | 5 | 5 | 5 | 27 |
| Sedaghatnezhad *et al.*, 2021<sup>29</sup> | 9 | 0 | 5 | 4 | 5 | 23 |
| Sled *et al.*, 2010<sup>36</sup> | 9 | 0 | 5 | 3 | 5 | 22 |
| Vincent and Vincent, 2020<sup>27</sup> | 10 | 1 | 6 | 5 | 5 | 27 |
| Zhu *et al.*, 2016<sup>25</sup> | 10 | 1 | 6 | 5 | 5 | 27 |

Total scores ranged from 19 to 29, indicating variation in methodological quality across the included studies. Strengths were generally observed in the reporting of study characteristics, interventions, outcome measures and statistical analyses. However, methodological limitations were identified, particularly regarding participant blinding, allocation concealment and external validity. The representativeness of study populations and settings was frequently unclear, potentially limiting the generalisability of the findings.

#### Changes in primary outcome measures

##### Spatiotemporal parameters

Changes in spatiotemporal gait parameters are presented in Figure 2.

**Figure 2.**
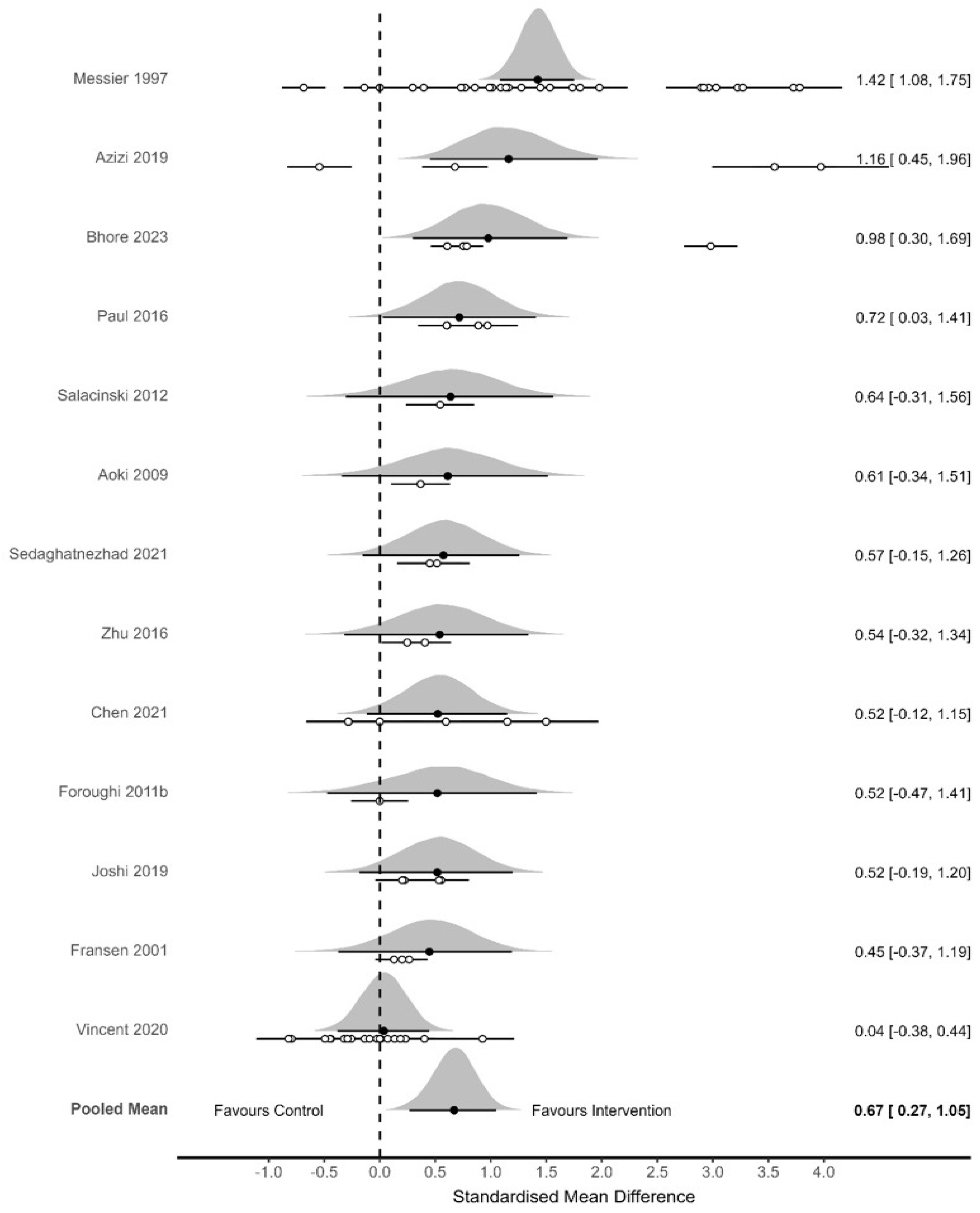
Forest plot for changes in spatiotemporal parameters (primary outcome). The black dots represent the standardised mean difference (SMD) effect size whilst the left and right extremes of the dots represent the corresponding 95% credible intervals. The overall SMD (pooled mean) with the corresponding credible intervals is also presented in this forest plot.

The initial meta-analysis conducted on all spatiotemporal outcomes (13 studies, 74 effect sizes and total N = 736) produced a pooled standardised mean difference effect size favouring physical activity intervention (SMD_0.5_ = 0.67 [95% CrI: 0.27 to 1.05]; between study SD *τ* = 0.56 [75% CrI: 0.39 to 0.80]; interclass correlation coefficient ICC = 0.53 [75% CrI: 0.09 to 0.77]).

The probability that the pooled mean effect size exceeded, zero, small, medium, and large effect sizes was >0.999, 0.978, 0.779, and 0.258, respectively. No meaningful changes, other than reduced precision, were identified when the analysis was split based on outcomes obtained over shorter (≤84 days; SMD_0.5_ = 0.79 [95% CrI: 0.46 to 1.1] or longer (>84 days; SMD_0.5_ = 0.62 [95% CrI: −0.36 to 1.5]; Figure 2) time periods.

Gait velocity was the most assessed spatiotemporal parameter [20, 24, 25, 27–34]. The cadence (the number of steps taken per minute) [20, 24, 26–28, 30, 32, 35], stride [20, 24, 26, 28–30, 32, 35] and step [24–27, 32, 35] lengths improved from baseline in studies that measured these parameters regardless of the exercise/physical activity intervention types.

Step width was significantly decreased following multi-component [35] and either concentrically or eccentrically focused resistance exercise interventions [27, 35], while two other studies did not identify improvement in this parameter assessed following the aquatic [26] or aerobic [28] exercise. Furthermore, studies found no significant difference in the swing and stance phase before and after the aerobic and resistance training programs [20, 28].

Overall, the results suggest that exercise has a moderate-to-large positive effect on gait timing and speed variables.

##### Kinematic parameters

Changes in kinematic gait parameters are presented in Figure 3.

**Figure 3.**
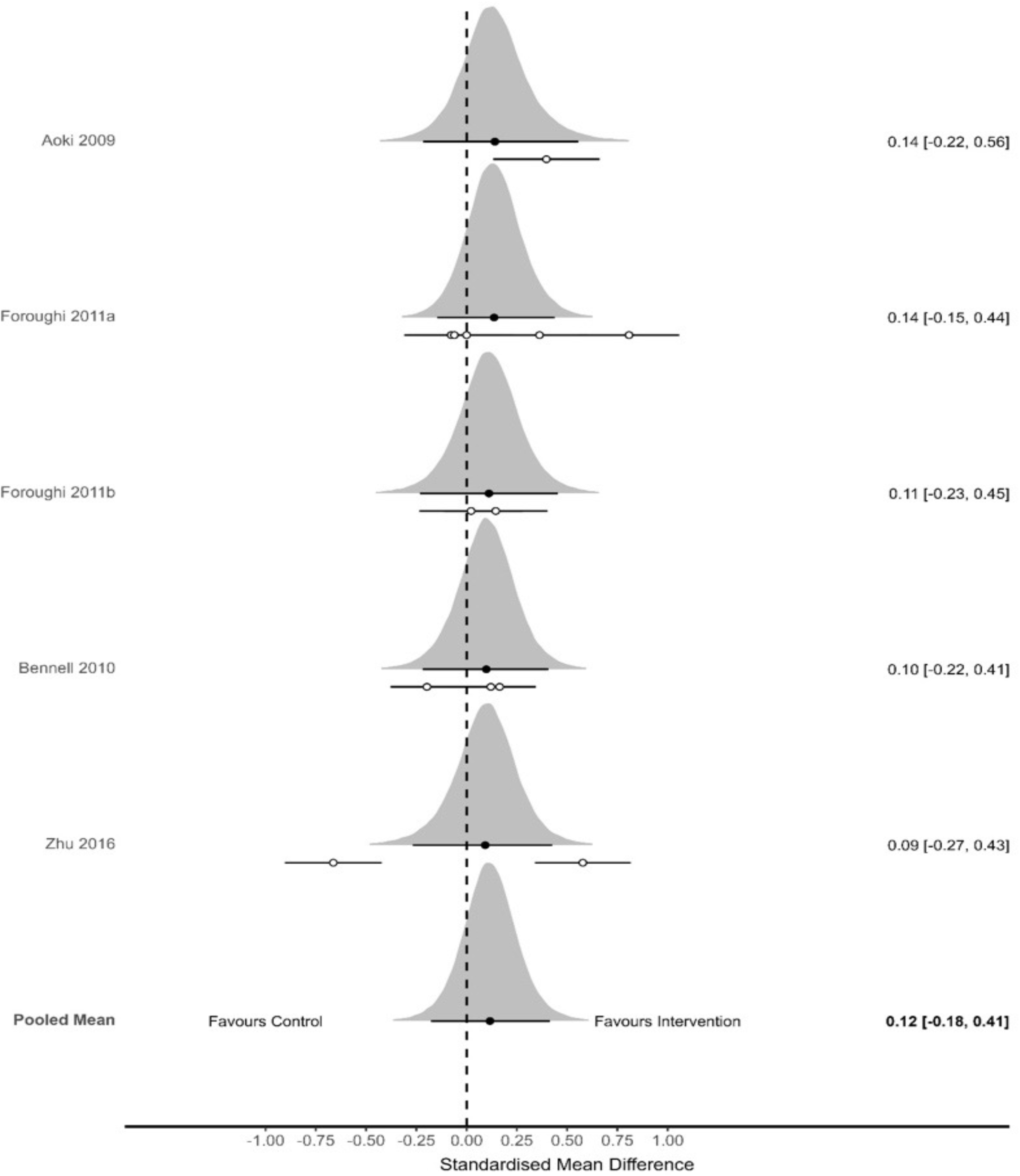
Forest plot for changes in kinematic parameters (primary outcome). The black dots represent the standardised mean difference (SMD) effect size whilst the left and right extremes of the dots represent the corresponding 95% credible intervals. The overall SMD (pooled mean) with the corresponding credible intervals is also presented in this forest plot.

The initial meta-analysis conducted on all kinematic outcomes (5 studies, 13 effect sizes and total N = 279) produced an uncertain pooled standardised mean difference with central distribution close to zero (SMD_0.5_ = 0.12 [95% CrI: −0.18 to 0.41]; *τ* = 0.08 [75% CrI: 0.01 to 0.24]; ICC = 0.85 [75% CrI: 0.21 to 0.99]; Figure 3).

The probability that the pooled mean effect size exceeded, zero, small, medium, and large effect sizes was 0.809, 0.259, 0.010, and <0.001, respectively. No meaningful changes, other than reduced precision, were identified when the analysis was split based on outcomes obtained over shorter (≤84 days; SMD_0.5_ = 0.13 [95% CrI: −0.96 to 1.3] or longer (>84 days; SMD_0.5_ = 0.11 [95% CrI: −0.40 to 0.61]) time periods.

Overall results suggest that exercise has a minimal effect on joint angles and movement patterns.

##### Kinetic parameters

Changes in kinetic gait parameters are presented in Figure 4.

**Figure 4.**
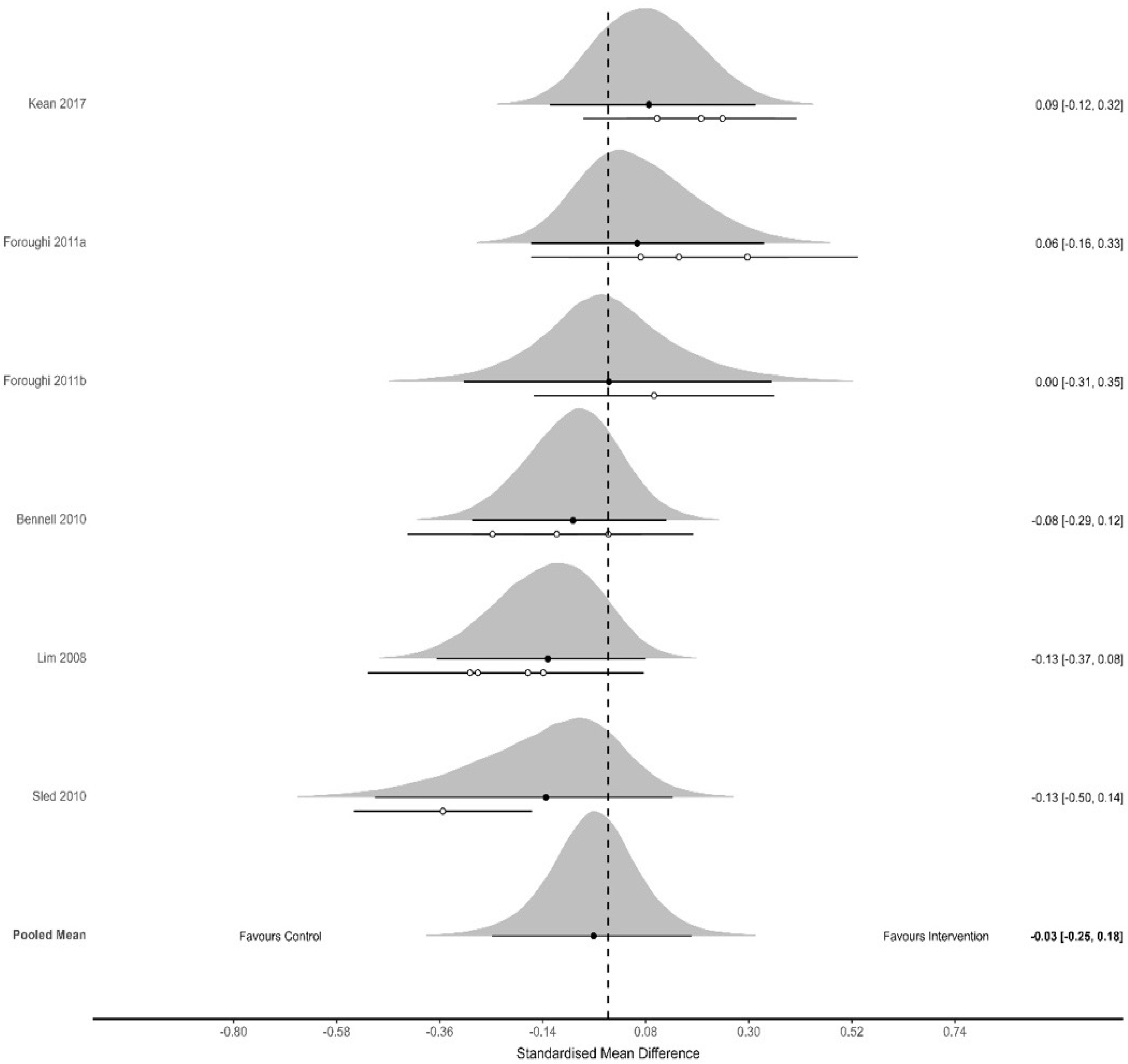
Forest plot for changes in kinetic parameters (primary outcome). The black dots represent the standardised mean difference (SMD) effect size whilst the left and right extremes of the dots represent the corresponding 95% credible intervals. The overall SMD (pooled mean) with the corresponding credible intervals is also presented in this forest plot.

The initial meta-analysis conducted on all kinetic outcomes (6 studies, 15 effect sizes and total N = 473) produced an uncertain pooled standardised mean difference (SMD_0.5_ = −0.03 [95% CrI: −0.25 to 0.18]; *τ* = 0.15 [75% CrI: 0.04 to 0.29]; ICC = 0.15 [75% CrI: 0.01 to 0.84]; Figure 4).

The probability that the pooled mean effect size exceeded, zero, small, medium, and large effect sizes was 0.369, 0.018, 0.001, and <0.001, respectively. Potential meaningful changes were identified when the analysis was split based on outcomes obtained over shorter (≤84 days; SMD_0.5_ = −0.09 [95% CrI: −0.41 to 0.19] but substantial uncertainty for longer (>84 days; SMD_0.5_ = 0.15 [95% CrI: −0.86 to 1.2]) time periods.

Overall results suggest that exercise does not reliably change joint-loading mechanics across studies.

#### Changes in secondary outcome measure

##### Pain

Changes in pain are presented in Figure 5.

**Figure 5.**
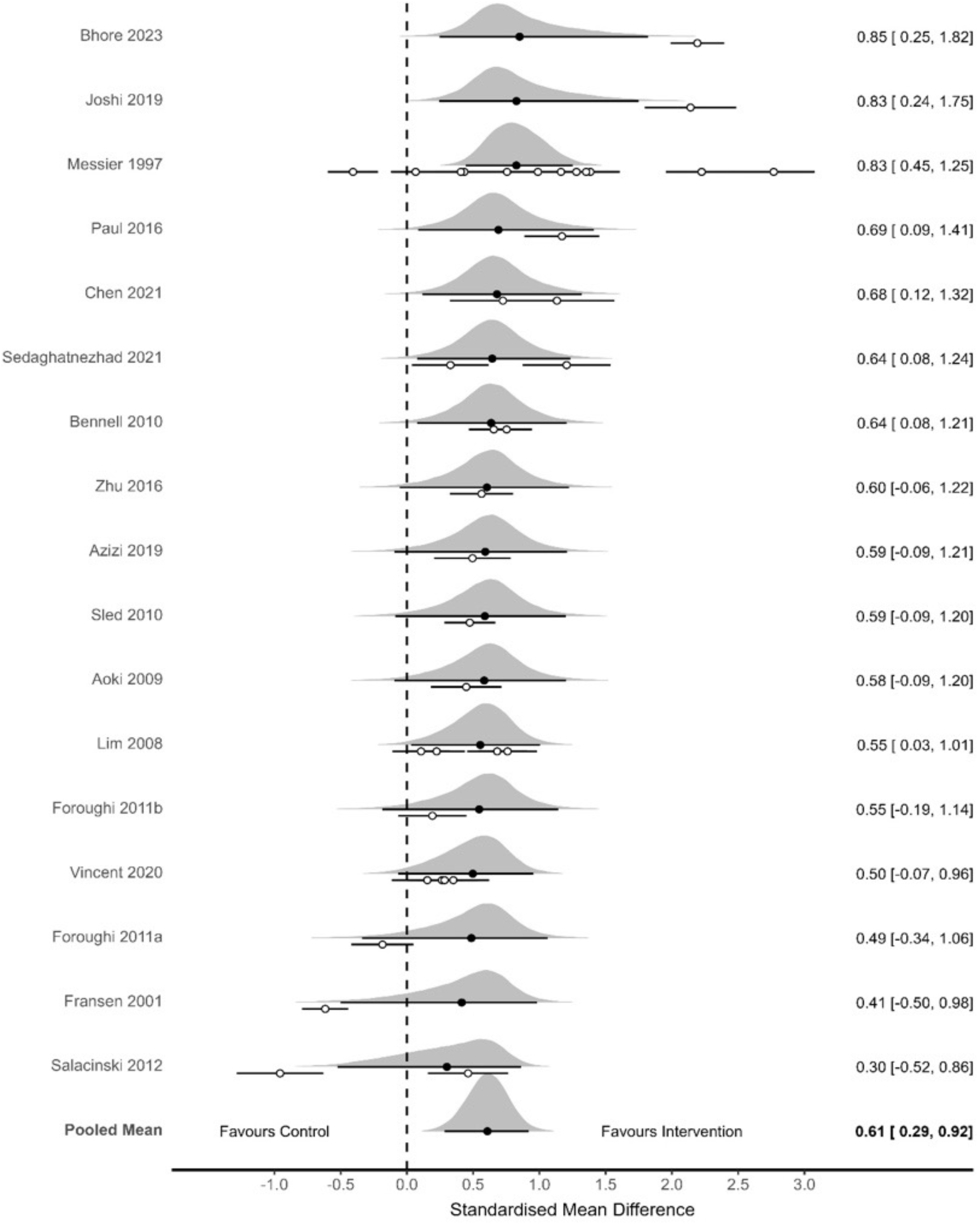
Forest plot for changes in pain (secondary outcome). The black dots represent the standardised mean difference (SMD) effect size whilst the left and right extremes of the dots represent the corresponding 95% credible intervals. The overall SMD (pooled mean) with the corresponding credible intervals is also presented in this forest plot.

The initial meta-analysis conducted on all pain outcomes (17 studies, 38 effect sizes and total N = 1102) produced a pooled standardised mean difference effect size favouring physical activity intervention (SMD_0.5_ = 0.61 [95% CrI: 0.29 to 0.92]; *τ* = 0.26 [75% CrI: 0.06 to 0.52]; ICC = 0.74 [75% CrI: 0.18 to 0.98]; Figure 5).

The probability that the pooled mean effect size exceeded, zero, small, medium, and large effect sizes was >0.999, 0.993, 0.757, and 0.109, respectively. No meaningful changes, other than reduced precision, were identified when the analysis was split based on outcomes obtained over shorter (≤84 days; SMD_0.5_ = 0.67 [95% CrI: 0.23 to 1.1] or longer (>84 days; SMD_0.5_ = 0.48 [95% CrI: −0.15 to 0.92]) time periods.

Studies revealed that muscle strengthening resistance [21–24, 30, 33, 36, 37], and aerobic [28, 34] exercise interventions reduced pain experienced by participants compared with study control groups. Overall results show moderate improvement in pain, sustained but reduced over time.

## Discussion

This systematic review and meta-analysis synthesised evidence from 18 studies to evaluate the effects of exercise and physical activity on gait biomechanics and pain in individuals with knee osteoarthritis. The principal finding is that exercise interventions produce moderate and clinically meaningful reductions in pain alongside consistent improvements in spatiotemporal gait parameters, while kinematic and kinetic adaptations at the knee joint appear minimal or uncertain. These results reinforce the role of exercise as a first line non-pharmacological strategy [39], but also highlight important biomechanical limitations that may inform the future intervention strategies.

The cellular mechanisms underlying pathogenesis of osteoarthritis are still not yet fully understood. Previous studies investigated molecular mechanisms underpinning articular cartilage degradation [40], while exercise and physical activity have been shown to play an important role in preserving synovial joint structural integrity by reducing inflammation and protecting articular cartilage [41].

The most robust effect observed was for pain reduction with a high probability of exceeding a small to moderate clinical threshold. This finding is consistent with OARSI recommendations that identify exercise as a core intervention for the management of knee osteoarthritis. This also aligns with existing clinical guidelines recommending exercise for symptom management in knee osteoarthritis and supports the hypothesis that both peripheral mechanisms including improvements in muscle strength, proprioception and joint stability, and central mechanisms such as exercise-induced pain modulation and alterations in pain processing, contribute to symptom relief [42].

The Bayesian analysis indicated approximately 76% probability that exercise produces at least a moderate reduction in pain. Notably multi-component and mind-body interventions appeared particularly effective, likely due to their ability to simultaneously target strength, flexibility, balance, and neuromuscular control while also addressing psychosocial aspects such as fear of movement.

Improvements in spatiotemporal gait parameters were also evident with consistent increases in walking speed, cadence, and step/stride length across studies. These changes suggest enhanced functional mobility and walking efficiency, which are clinically meaningful since the gait speed is an established indicator of physical function, disability status, and health-related quality of life in individuals with knee osteoarthritis [43]. The uniformity of these findings across diverse exercise modalities indicates that general increases in physical capacity, rather than modality-specific adaptations, may underpin these improvements.

In contrast, kinematic and kinetic outcomes showed negligible or uncertain effects. These findings suggest that while patients walk faster and more efficiently following intervention, the underlying joint mechanics, particularly those linked to disease progression, such as knee adduction moment, remain largely unchanged. This discrepancy may reflect the complexity of modifying joint loading patterns which are influenced not only by muscle strength but also by structural alignment, joint morphology, and long-standing motor patterns.

Altered gait biomechanics in knee osteoarthritis are characterised by increased knee adduction moments, reduced knee flexion angles and moments during stance, altered hip kinetics, and progressive reductions in lower-limb range of motion as disease severity increases [4, 5]. Furthermore, individuals with knee osteoarthritis commonly exhibit deficits in hip and knee muscle strength, reduced knee extension during gait, and altered frontal-plane mechanics associated with disease severity [44, 45].

The present findings are consistent with recent systematic reviews demonstrating that exercise may improve pain and physical function in knee osteoarthritis while having limited effects on joint loading metrics such as the knee adduction moment [46, 47] suggesting that traditional strengthening programs alone may be insufficient to alter medial knee joint loading. Unlike previous reviews that primarily focused on pain and function or biomechanics only, the present review examined gait biomechanics, distinguished between spatiotemporal, kinematic and kinetic domains and integrated biomechanical and probabilistic synthesis.

Emerging literature suggests that targeted biomechanical interventions, such as gait retraining [48, 49], valgus bracing [50], or neuromuscular re-education [46], may be required to meaningfully influence these parameters. The modest improvements in some of the kinematic variables such as pelvic control or range of motion observed in individual studies further support the idea that specific, targeted interventions, but not generic exercise programs, are necessary to modify joint mechanics.

These findings have several implications for clinical practice. Exercise such as multi-component and mind-body approaches may be recommended for pain reduction and functional improvement. Improvements in walking speed and related parameters may serve as practical and sensitive clinical indicators of progress. Furthermore, to influence disease progression, clinicians may need to combine exercise with biomechanical interventions such as gait retraining and orthotics. The limited changes in kinetic variables suggest that exercise alone may not substantially modify some biomechanical factors proposed to be associated with disease progression. Programs should be targeted and individualised, considering patient-specific factors such as alignment, severity, and functional limitations.

## Data Availability

All data analysed in this systematic review were obtained from previously published studies. The data supporting the findings of this review are presented in the manuscript and its supplementary materials. No new individual-level participant data were generated or collected.

## Acknowledgements and affiliations

This work was supported by the Anglia Ruskin University (ARU) Vice Chancellor’s Scholarship (to HJS). The findings were previously presented orally and as posters at conferences of the International Society of Biomechanics (ISB), the European Calcified Tissue Society (ECTS) and the British Orthopaedic Research Society (BORS).

The ECTS 2026 Congress abstract was published in *JBMR Plus* as part of the *Abstracts of the ECTS 2026 Congress* supplement (2026;10(Suppl 1):i1–i319; doi:10.1093/jbmrpl/ziag066).

