## Supplemental Table 3 for "The effects of exercise and physical activity on gait biomechanics and pain in knee osteoarthritis: A systematic review and meta-analysis"

**Table S3. Search strategy**

| **Condition** | **Joint/anatomical site** | **Physical activity or exercise** | **Biomechanical data** | **Boolean operators** | **Grouping** | **Truncation** | **Proximity operators** |
| --- | --- | --- | --- | --- | --- | --- | --- |
| Osteoarthritis | Knee | Exercise* | gait | AND, OR | parenthesis ( ) | asterisk * | PubMed  [tiab:~4] |
| Knee osteoarthritis |  | Physical activity | kinemat* |  |  |  | Web of Science  NEAR/4 |
| Osteoarthritis, Knee |  | Physical activities | kinet* |  |  |  | SPORTDiscus  N/4 |
| Osteoarthritic knee |  | Sport exercise |  |  |  |  | Scopus  W/4 |
|  |  | Physical exercise |  |  |  |  |  |
