## Supplemental Table 4 for "The effects of exercise and physical activity on gait biomechanics and pain in knee osteoarthritis: A systematic review and meta-analysis"

**Table S4. Search results**

| **Database** | **Search strategy** | **Filters applied** | **Number of articles** |
| --- | --- | --- | --- |
| **PubMed** | (("Osteoarthritis, Knee" OR "knee osteoarthritis") OR ("knee osteoarthritis" OR "osteoarthritic knee"[tiab :~4]) AND (Gait OR Kinemat* OR Kinet*) AND (Exercise*OR Physical activity OR Physical activities OR Physical Exercise OR Sport Exercise)) | Human (MeSH) | 1026 |
| **SPORTDiscus** | (("Osteoarthritis, Knee" OR "Knee osteoarthritis") OR ("knee n/4 osteoarthritis" OR osteoarthritic n/4 knee") AND (Gait OR Kinemat* OR Kinet*) AND (Exercise* OR Physical Activity OR Physical Activities OR Physical Exercise OR Sport Exercise)) | None | 143 |
| **Web of Science** | (("Osteoarthritis, Knee" OR "Knee osteoarthritis") OR ("knee near/4 osteoarthritis" OR osteoarthritic near/4 knee") AND (Gait OR Kinemat*OR Kinet*) AND (Exercise* OR Physical Activity OR Physical Activities OR Physical Exercise OR Sport Exercise)) | None; human studies could not be filtered | 747 |
| **Scopus** | (({Osteoarthritis, Knee} OR {knee osteoarthritis}) OR ("knee w/4 osteoarthritis" OR "osteoarthritic w/4 knee") AND (gait OR kinemat* OR kinet*) AND (exercise* OR "Physical activity" OR "Physical activities" OR "Physical Exercise" OR "Sport Exercise")) | None | 686 |
