## Supplemental Table 5 for "The effects of exercise and physical activity on gait biomechanics and pain in knee osteoarthritis: A systematic review and meta-analysis"

**Table S5. Codebook**

| **Column** | | **Heading** | **Description** |
| --- | --- | --- | --- |
| **Study Details** | A | Initials Reviewer | Identification of individual extracting information |
|  | B | Study Number | The number assigned to the study |
|  | C | Covidence Identifier | Reference number for Covidence |
|  | D | Author | First author surname *et al.* |
|  | E | Year | Year of publication |
|  | F | Title | Study title |
|  | G | Country | Country where study was conducted |
|  | H | Journal | Journal name |
|  | I | Aims/Purpose | Study aims/purpose |
|  | J | Knee Diagnosis | 1 = Symptomatic; 2 = Kellen-Lawance 1-2; 3 = Kellen-Lawrance 3-4; 4 = ACR Scale 1-2; 5 = ACR Scale 3-4; 6 = Medial; 7 = Bilateral; 8 = Grade 1 to 3 (No classification criteria); 9 = No classification/No information given; 10 = ACR (No grade given); 11=Outerbridge 1-4 |
|  | K | Study Design | 1 = RCT; 2 = Quasi-experimental; 3 = Comparative or experimental |
|  | L | Age Mean | Mean age of study sample as a whole |
|  | M | Age SD | Standard deviation age of study sample as a whole |
|  | N | Baseline Total N | Total sample across all interventions measured at baseline |
|  | O | Exercise Description | Brief description of exercise of a study as a whole |
|  | P | Exercise Description | Brief description of exercise highlighting muscles and lower limb joints involved |
|  | Q | Sex (% Female) | Percentage female of study sample as a whole |
|  | R | BMI Mean | Mean BMI of study sample as a whole |
|  | S | BMI SD | Standard deviation of BMI of study sample as a whole |
|  | T | Symptom Duration Mean (Months) | Mean symptom duration reported in months |
|  | U | Symptom Duration SD (Months) | Standard deviation symptom duration reported in months |
|  | V | Population Comments | Any additional information relevant to the participants investigated including diagnostic criteria |
| **Outcomes** | W | Outcome Category - Gait Characteristics | 1 = Kinematic (KM); 2 = Kinetic (KT); 3 = Spatiotemporal (ST) |
|  | X | Kinematic Parameters (KM) | 1 = G-ROM; 2 = Initial contact angle of knee joint; 3 = Maximal angle of knee joint; 4 = Shank adduction angle; 5 = Knee adduction angle; 6 = KFBWH; 7 = KEBWH; 8 = HEBWT; 9 = HFBWT; 10 = Contralateral pelvic drop; 11 = Maximum ipsilateral trunk lean; 12 = Toe-out stance |
|  | Y | Kinetic Parameters (KT) | 1 = KAM impulse (area under curve); 2 = PKAM; 3 = Max Rate of Loading; 4 = Peak early stance (fZ); 5 = Ave Loading Rate; 6 = KAM Peak 1; 7 = KAM Peak 2; 8 = HAM Peak 1; 9 = HAM Peak 2 |
|  | Z | Spatiotemporal (ST) | 1 = Step length; 2 = Step/stride width; 3 = Stride length; 4 = Cadence; 5 = Velocity (walking speed/walking velocity/speed); 6 = Swing phase; 7 = Stance phase; 8 = Double limb support; 9 = Stride Time (Coeff of Variance); 10 = Swing Time (Coeff of Variance); 11 = single leg support; 12 = Velocity Coeff of Variance; 13 = Stride Length (Coeff of Variance); 14 = Double limb support (Coeff of Variance) |
|  | AA | Outcome Tool (Gait) | Description of outcome tool(s) |
|  | AB | Outcome Category - Pain | 1 = WOMAC; 2 = VAS; 3 = KOOS; 4 = NRS; 5 = Other scale (referenced) |
|  | AC | Pain Outcome Subscales | Subscales used for pain |
|  | AD | Measurement Time (Days) | Time of measurement in days |
|  | AE | First Follow up | Time of first follow up in weeks if applicable |
|  | AF | Second Follow up | Time of the second follow up in weeks if applicable |
|  | AG | Third Follow up | Time of the third follow up in weeks if applicable |
|  | AH | Outcome | Specific outcome measured |
|  | AI | Outcome Domain | The outcome domain measured |
|  | AJ | Reflection | 1 = Increase in outcome indicates positive treatment; -1 = Decrease in outcome indicates positive treatment |
| **Intervention** | AK | Exercise/Physical Activity Type | 1 = Resistance; 2 = Strength; 3 = Vibration; 4 = Aquatic; 5 = Open and closed kinetic chain; 6 = Stretching; 7 = Mind Body; 8= NMT; 9 = Retro walking; 10 = Walking/Basic exercise; 11 = Aerobic; 12 = Multicomponent |
|  | AL | Intervention N (Group 1) | Intervention sample size at specified time |
|  | AM | Intervention N (Group 2) | Intervention sample size at specified time |
|  | AN | Intervention Total Duration (days) | Total duration of exercise intervention in days |
|  | AO | Intervention Adherence % | Reporting of adherence to exercise (reported as a percentage) if applicable |
|  | AP | Intervention Location | Location exercise/physical activity was performed  1 = Home; 2 = Clinic; 3 = Fitness facility; 4 = NR; 5 = N/A |
|  | AQ | Intervention Volume | Numerical value describing volume |
|  | AR | Intervention Volume Category | 1 = Duration of session (mins); 2 = sets * repetitions; 3 = number of repetitions; 4 = number of sets |
|  | AS | Intervention Volume Comments | Any additional information relevant |
|  | AT | Intervention Intensity | Numerical value describing intensity |
|  | AU | Intervention Intensity Category | 1 = Absolute; 2 = Relative |
|  | AV | Intervention Frequency | Number of sessions per week. Where there is progression, average value is to be entered |
|  | AW | Intervention Frequency Comments | Any additional information relevant |
|  | AX | Intervention Progression | Multiple types to be selected as required  1 = No progression; 2 = NR; 3 = Progression volume; 4 = Progression intensity; 5 = Progression frequency; 6 = Progression specificity; 7 = Progression capacity; 8 = Other |
|  | AY | Intervention Progression Comments | Any additional information relevant |
| **Control** | AZ | Control Comparator | Description of the control group |
|  | BA | Control N | The control sample size |
|  | BB | Control Comparator Comments | Any additional information relevant |
| **Data** | BC | Control Baseline Age (Mean) | Baseline mean age for control |
|  | BD | Control Baseline Age (SD) | Baseline standard deviation age for control |
|  | BE | Control Baseline BMI (Mean) | Baseline mean BMI for control |
|  | BF | Control Baseline BMI (SD) | Baseline standard deviation BMI for control |
|  | BG | Intervention Baseline Age (Mean) | Baseline mean age for intervention |
|  | BH | Intervention Baseline Age (SD) | Baseline standard deviation age for intervention |
|  | BI | Intervention Baseline BMI (Mean) | Baseline mean BMI for intervention |
|  | BJ | Intervention Baseline BMI (SD) | Baseline standard deviation BMI for intervention |
|  | BK | Measurement Comments | State if a different value has been entered for means (e.g. median), a different value for standard deviations (e.g. standard error, IQR, percentiles, distance from mean to upper bound). Provide the relevant statistic (width of CI’s, width of percentiles).  Also state if data has been extracted by digitisation |
|  | BL | Control Measurement Mean | Mean of outcome for control at stated time point |
|  | BM | Control Measurement SD | Standard deviation of outcome for control at stated time point |
|  | BN | Control Measurement CI (lower) | Confidence Interval (lower) of outcome for control at stated time point |
|  | BO | Control Measurement CI (upper) | Confidence Interval (upper) of outcome for control at stated time point |
|  | BP | Control Measurement Post-intervention Mean | Mean of outcome for control post-intervention |
|  | BQ | Control Measurement Post-intervention SD | Standard deviation of outcome for control post-intervention |
|  | BR | Control Measurement Post-intervention CI (lower) | Confidence Interval (lower) of outcome for control post-intervention |
|  | BS | Control Measurement Post-intervention CI (upper) | Confidence Interval (upper) of outcome for control post-intervention |
|  | BT | Intervention Measurement Mean | Mean of outcome for intervention at stated time point |
|  | BU | Intervention Measurement SD | Standard deviation of outcome for intervention at stated time point |
|  | BV | Intervention Measurement CI (lower) | Confidence Interval (lower) of outcome for intervention at stated time point |
|  | BW | Intervention Measurement CI (upper) | Confidence Interval (upper) of outcome for intervention at stated time point |
|  | BX | Intervention Measurement Post-intervention Mean | Mean of outcome post-intervention |
|  | BY | Intervention Measurement Post-intervention SD | Standard deviation of outcome post-intervention |
|  | BZ | Control Measurement Post-intervention CI (lower) | Confidence Interval (lower) of outcome post-intervention |
|  | CA | Control Measurement Post-intervention CI (upper) | Confidence Interval (upper) of outcome post-intervention |

*** Outcome Specific**
